# Analysis of recurrent research pathways for assessing and improving effectiveness in life sciences laboratories

**DOI:** 10.1101/2023.01.09.23284360

**Authors:** E. Andrew Balas, Charmi Patel, Ben Ewing, Nauka Patel, Tiana Curry, Scott Wise, Yara H Abdelgawad

## Abstract

**Background:** Life sciences research often turns out to be ineffective. Our aim was to develop a method for mapping repetitive research processes, detecting practice variations, and exploring inefficiencies.

**Methods:** Three samples of R&I projects were used: companion diagnostics of cancer treatments, identification of COVID-19 variants, and COVID-19 vaccine development. Major steps involved: defined starting points, desired end points; measurement of transition times and success rates; exploration of variations, and recommendations for improved efficiency.

**Results:** Over 50% of CDX developments failed to reach market simultaneously with new drugs. There were significant variations among phases of co-development (Bartlett test P<0.001). Length of time in vaccine development also shows variations (P<0.0001). Similarly, subject participation indicates unexplained variations in trials (Phase I: 489.7 (±461.8); Phase II: 857.3 (±450.1); Phase III: 35402 (±18079).

**Conclusion:** Analysis of repetitive research processes can highlight inefficiencies and show ways to improve quality and productivity in life sciences.

---

Life sciences research has produced many landmark discoveries for public health improvement but many research projects turned out to be ineffective. Begley and Prinz estimated that the amount of non-reproducible research is somewhere between 75 to 89%, astoundingly high rates (Begley, 2012, Prinz, 2011). Improving the quality and productivity of scientific research is a paramount societal interest. Part of the difficulty relates to the complexity of the process involved in translating new biologic and technologic developments into routine clinical medicine (Parkinson, 2012).

Scientific research is inherently creative, variable, and unpredictable, making it difficult to help with quality improvement efforts. On the other hand, several important research processes turn out to be repetitive and therefore analyzable for improving performance. In such cases, information can be collected about multiple occurrences of the same or similar processes and analyzed for optimization. The following three examples from life sciences research illustrate the many repetitive processes and the need for improvement:

A growing number of companion diagnostics aims to identify patients that will respond to targeted therapies, thus increasing efficacy and safety of the drug. The pace of progress in linking targeted therapies with appropriately characterized biomarkers and patients is known to be frustratingly slow. The post launch delay is approximately 4.5 years, and it takes from 1.5 to 5 years to offer the test by labs to achieve optimal rate after treatment introduction (Keeling et al, 2020). Failure to obtain the right target for the cancer treatment led to the failure to get the desired results in many drugs clinical trials which has overall impact on initial stages of co-development process (Hu, 2019). Failure to obtain information about different diagnostic metrics affects the evaluation of clinical validity of the assay (Jorgensen, 2018).

Virus variants are coming up quickly and randomly, but we are slow in identifying them. The current process to identify and classify SARS-COV-2 variants is a detailed process that takes up to a month from the time of a positive tests for COVID-19 to the identification and classification of a variant. State health departments prepare and send positive COVID-19 samples to the CDC weekly or biweekly. It then takes about ten days for the CDC or their contracted agencies to perform genetic sequencing and identify variants (CDC, 2021). The US is behind 30 other countries in the amount of sequencing done during the pandemic despite having the equipment and experience to do more (Maxmen, 2021). Much of COVID-19 testing is done in labs that do not have genomic sequencing capabilities, therefore samples are often discarded due to the need for extra storage and labor needed to store samples (Maxmen, 2021).

The rapid spread and devastating consequences of the COVID-19 pandemic have changed the vaccine development processes. In response to the need to expedite development which previously took many years, researchers pushed for adjustments to regulation and processes, such as the decision to begin Phase 1 clinical trials before the completion of animal testing (Nguyen, 2021). The Pfizer and Moderna vaccines’ Phase 1 and preclinical animal trials were initiated in parallel as early as April 2020, with parallel Phases 2 and 3 being run three months later (Kieny, 2021). In early 2020, it was unethical to use human challenge studies for SARS-CoV-2 vaccine development (Kahn, 2020). This made it difficult to determine vaccine efficacy in variant subjects and this has been problematic as the controlled human infection models can accelerate vaccine development (Rahman, 2019).

In response to these challenges, comparative study and reengineering of research processes may help to develop faster research response systems. The purpose of this study was to develop and pilot test the systematic analysis of repetitive research processes and find what appears to be ineffective or ready for improvement.

## Methods

The evaluation method of this study is rooted in data science that involves facts and statistics that are available from large databases or published literature. Analysis of recurring research involves the following steps: take a sample of R&I projects; clearly define starting point and define the desired endpoint; measure transition times and success rates; explore variations and weaknesses; and develop recommendation for improved efficiency.

### 1. Developing eligibility criteria of a repetitive research process

The first step is the identification of recurring research processes which happen with sufficient frequency to observe and analyze a sample of them. These selected research processes should have clearly identifiable starting and end points.

### 2. Sampling of R&I projects

After setting the eligibility criteria, the next step is to collect a sufficiently large sample research and development projects. It is desirable to have at least 10-30 randomly selected research and development processes to make statistical analysis sufficiently substantial.

### 3. Defining starting and desired end point

Such definitions are essential to set the boundaries of analysis and create opportunities for comparing multiple research processes and explore weaknesses or deficiencies. Typically, the process starts with the recognition of the need to launch a research process and ends when completion of the research delivers value that is universally recognizable (e.g., FDA approval of safety and efficacy)

### 4. Flowcharting the process of research

Visual presentation of either the actual or the ideal research process can be very valuable in identifying weaknesses and discrepancies. In much translational type research, the process includes discovery of the opportunity, preclinical study, clinical study with phase I, II and III clinical trials and finally FDA approval.

The flowchart should help to describe identifiable start and end times of all phases plus the identifiable intermediate results of each phase. The flow chart for the research process should support the assessment of the entire series of interconnected activities. For example, thin arrows can indicate the flow of the process, and bold arrows can highlight the difference between phases.

### 5. Collecting data about transition times and success rates

Independent variables include availability of the resources from the previous phase and the dependent variables include time and frequency. The data sources of this study included: FDA.gov, clinicaltrials.gov, PubMed. Google Scholar, CDC, USPTO, and others to extract relevant information. Research and commercial R&D database information about initiatives were also retrieved to study patterns of development and repurposing.

### 6. Exploring variations and weaknesses

Each or most flowcharted steps were evaluated for their transition time from the beginning to the end and for their success rates. Additionally, overall time spans were also evaluated from the beginning to the completion of the entire research process.

Subsequently, averages, differences, and standard deviations were calculated between the start of phases and for the overall development time. We used Bartlett’s test for the null hypothesis that all population variances are equal against the alternative that at least two are different.

### 7. Developing recommendation for improved efficiency

Based on the above-described information collection, unexplained variations and avoidable failure rates can be observed and recommended for improvement.

Suggested improvements in future research and development processes should offer higher quality results in a timely manner for better public health outcomes.

## Results

This study applied the above described research pathway assessment methodology and looked at the current routine processes in the (i) development of 14 drug-companion diagnostic test pairs as shown in) Table 1; (ii)development of 6 vaccines for the SARS-COV-2 virus; and (iii) detection and classification of 11 known variants to the SARS-COV-2 virus as of December 2021 (Table 1c). The estimated percent increase in transmission from the original SARS-COV-2 strain to 5 of the known variants is also displayed. The table also shows where and when the variants were first identified; the highest classification the variant received and when it received, and the current classification.

**Table 1a:** The development process of analyzed companion diagnostics pairs

| **Companion Diagnostics** | **Drugs** | **Biomarkers** | **Diseases** | **Was approved together?** | **Aproval of CDX** | **Aproval of drug** |
| --- | --- | --- | --- | --- | --- | --- |
| Abott Real time IDH1 | Tibsovo | IDH1 | Acute Myeloid Leukemia | Yes | Jul-18 | Jul-18 |
| FoundationOne CDx | Alecensa | ALK rearrangements | Non-small cell lung cancer | No | 17-Nov | Dec-15 |
| MRDx BCR-ABL test | Tasigna | Philadelphia Chromosome positive | Chronic Myeloid Leukemia | No | 17-Dec | Oct-07 |
| Cobas KRAS mutation test | Vectibix | KRAS mutation Negative | Colorectal cancer | No | May-15 | Sep-06 |
| BRCA analysis CDx | Lynparza | BRCA 1/2, HR+ AND HER2 Negative | Breast Cancer | Yes | Jan-18 | Jan-18 |
| Myriad mychoice CDx | Zejula | BRCA1/2 | Ovarian Cancer | No | Oct-19 | Mar-17 |
| BRCA analysis CDx | Lynparza | germline BRCA | Pancreatic Cancer | Yes | Dec-19 | Dec-19 |
| FoundationOne CDx | Erleada | mCRPC with BRCA1/2 | Prostate Cancer | No | Nov-17 | Feb-18 |
| HER2 FISH pharmDx | Herceptin (ENHERTU) | HER2 | Gastric or Gastroesophageal Cancer | N/A | Oct-10 | Jan-21 |
| Vysis CLL FISH Probe | Venetoclax | LSI TP53 | B-cell chronic lymphocytic leukemia | Yes | Apr-16 | Apr-16 |
| THXID BRAF assay | Tafinlar | BRAF | Melanoma | Yes | May-13 | May-13 |
| FoundationOne CDx | Pemazyre | FGFR2 | Cholangiocarcinoma | N/A | Nov-17 | Apr-20 |
| Therascreen FGFR RGQ RT-PCR test | Balversa | FGFR3 | Urothelial (Bladder) cancer | Yes | Apr-19 | Apr-19 |
| VENTANA MMR RXDX Panel | Jemperli | DMMR | Endometrial Carcinoma | Yes | Apr-21 | Apr-21 |

**Table 1b:** Length of time comparisons of phases in vaccine development projects (days).

| **Development of Vaccine** | **Diff. Preclinical-Phase 1** | **Diff. Phase I-Phase II** | **Diff. Phase II-Phase III** | **Diff. Phase III-FDA approval** |
| --- | --- | --- | --- | --- |
| BioNTech/Pfizer | 30.00 | 30.00 | 30.00 | 30.00 |
| Moderna | 22.00 | 73.00 | 58.00 | 0.00 |
| AstraZeneca | 26.00 | 28.00 | 97.00 | 122.00 |
| Janssen (Johnson & Johnson) | 105.00 | 0.00 | 66.00 | 156.00 |
| Novavax | 25.00 | 0.00 | - | - |
| GSK/Sanofi | 21.00 | 716.00 | 95.00 | - |
| Average (±SD) | 38.17 (±32.90) | 141.2 (±282.9) | 69.2 (±27.9) | 77.00 (±73.9) |

**Table 1c:** COVID-19 variant transmissibility, identification, and classification

| **Variants** | **Where First Identified** | **Earliest Documented Samples** | **Highest Classification** | **Current Variant Classification** | **Estimated Transmissibility increase (%)** |
| --- | --- | --- | --- | --- | --- |
| Alpha B.1.1.7 | United Kingdom | Sep-20 | VOC: December 29, 2020 | VBM | 29 |
| Beta B.1.351 | South Africa | May-20 | VOC: December 29, 2020 | VBM | 25 |
| Gamma P.1 | Japan/Brazil | Nov-20 | VOC: December 29, 2020 | VBM | 38 |
| Delta B.1.617.2 | India | Oct-20 | VOC | VOC | 97 |
| Kappa B.1.617.1 | India | Oct-20 | VOI: May 7, 2021 | VBM | 48 |
| Mean |  |  |  |  | 47.4 |
| Standard Deviation |  |  |  |  | 29.11 |
| Eta B.1.525 | Nigeria | Dec-20 | VOI: February 26,2021 | VBM |  |
| Omicron B.1.1.529 | Multiple countries | Nov-21 | VOC: November 26,2021 | VOC |  |
| Iota B.1.526 | New York, U.S. | Nov-20 | VOI: February 26,2021 | VBM |  |
| Mu B.1.621 | Columbia | Jan-21 | VBM: September 21, 2021 | VBM |  |
| Epsilon B.1.427 and B.1.429 | California, U.S | Sep-20 | VOC: March 19,2021 | VBM |  |
| Zeta P.2 | Brazil | Jan-21 | VOI: February 26, 2021 | VBM |  |

In addition to the table version of the research flow, results of the research process exploration and visualization can also be seen in three flowcharts. The process in the development of companion diagnostics can be seen in Figure 1a. Development of vaccines against Covid-19 are presented on Figure 1b. Finally, the process of variant identification of the SARS-COV-2 virus is shown in Figure 1c.

**Figure 1a:**
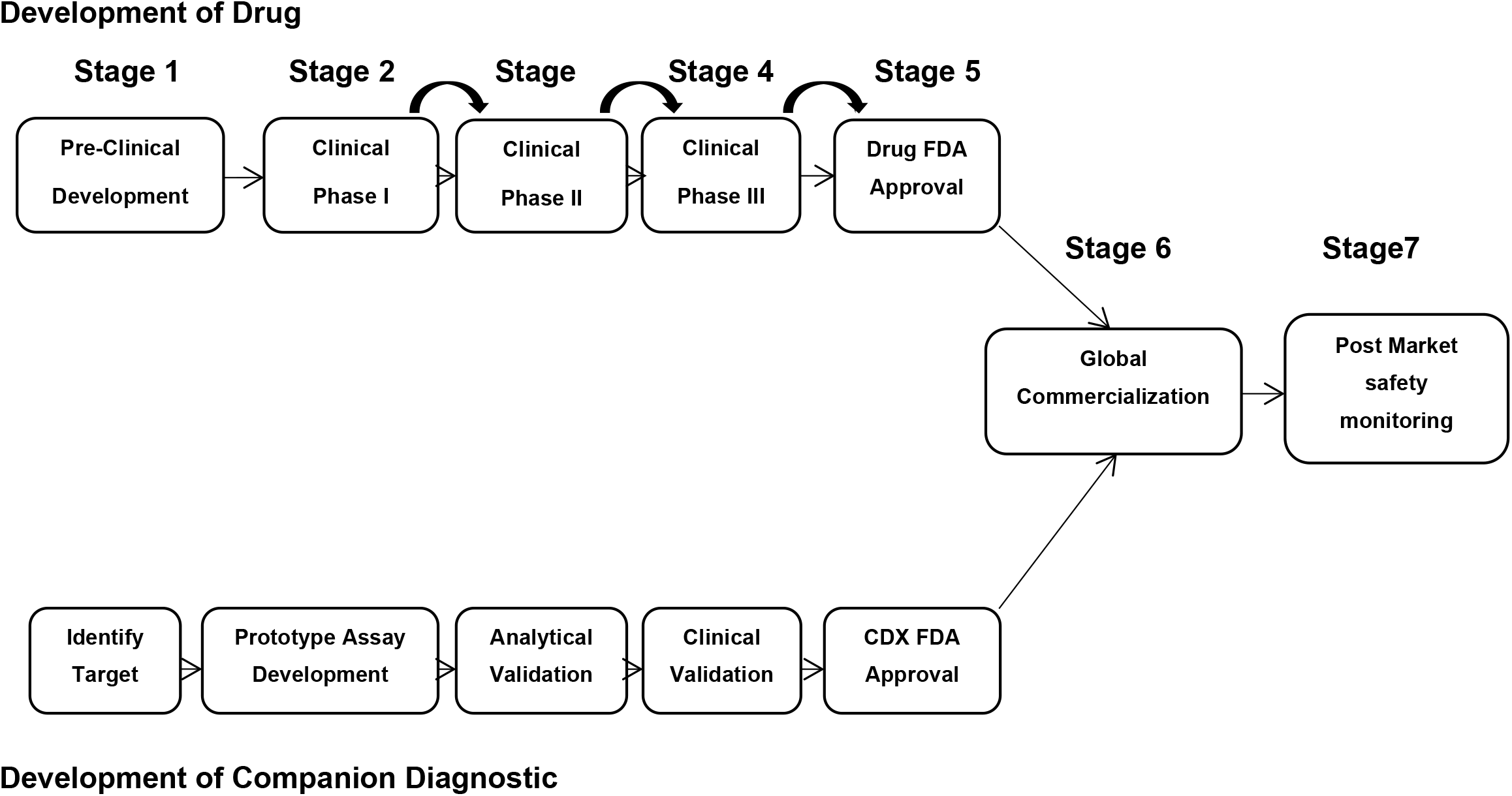
Ideal co-development process of drug and companion diagnostics

**Figure 1b:**
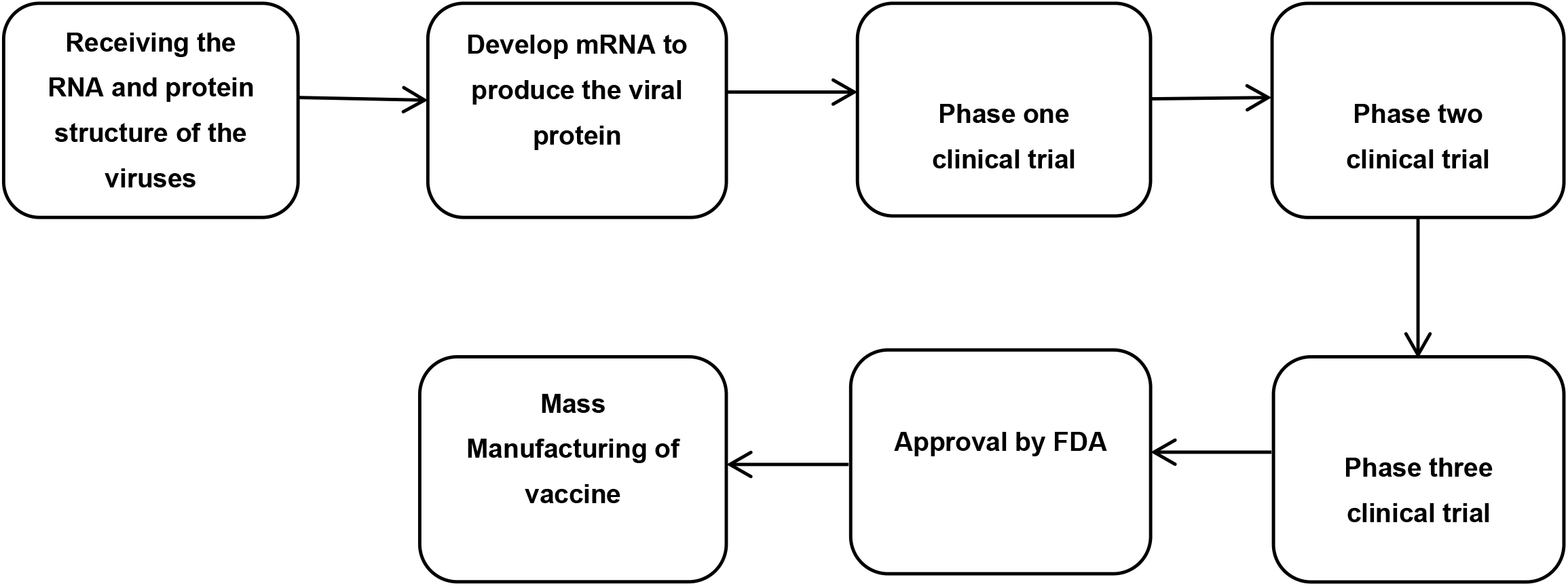
Current Process of Vaccine Development for COVID-19

**Figure 1c:**
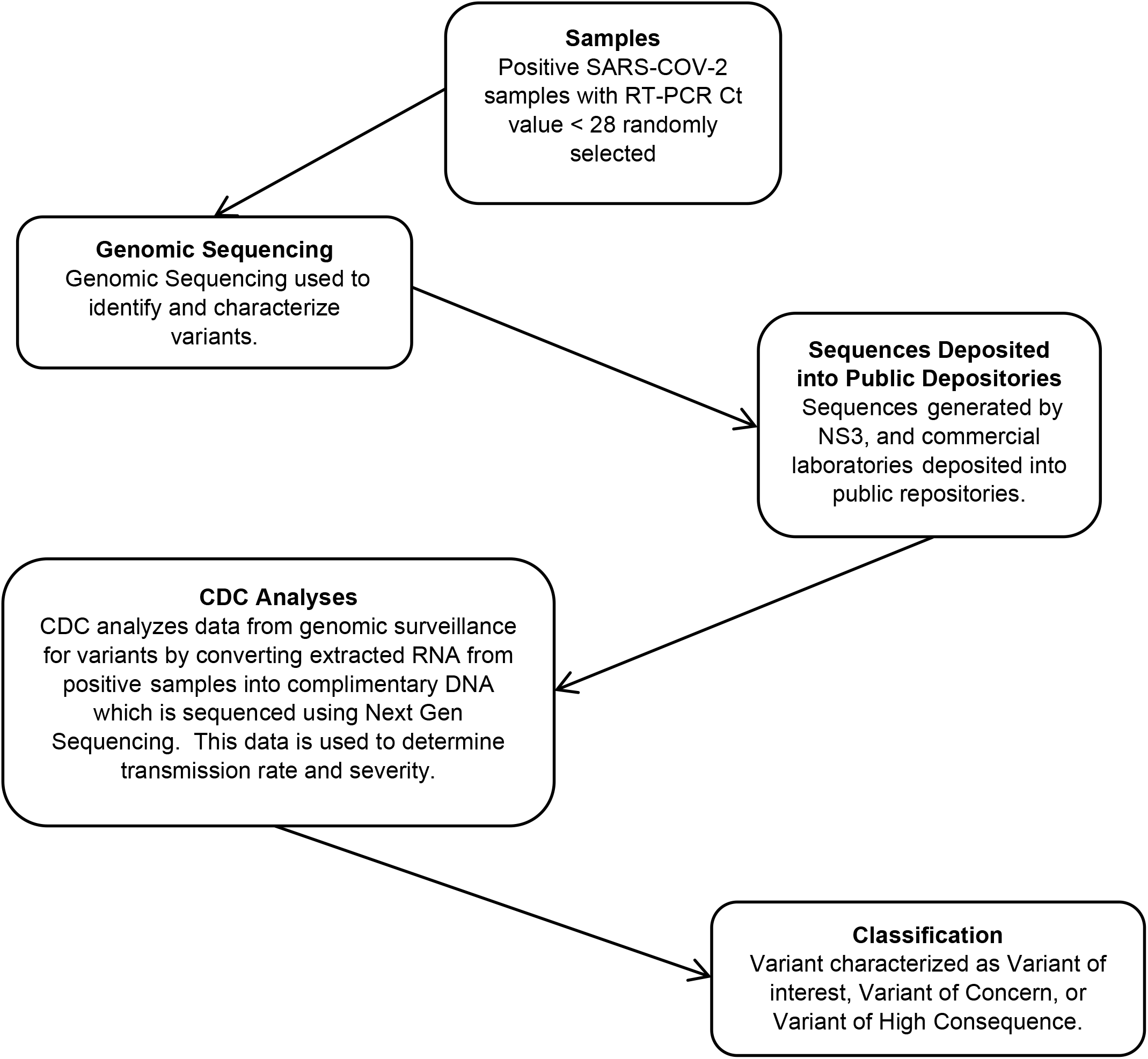
Current process of Identifying and Classifying Variants of SARs-COV-2

### Weaknesses of Transition Times

In the development of companion diagnostics, the average development time of the new drug and CDX is 5.12 and 4.72 years, respectively, but the 4.92 years approval gap is remarkable. Life-threatening side effects have also been detected with an average of 45.17% (± 19.98%) in phase 1 trials.

Obviously, timelines for the development of the mRNA-based Pfizer and Moderna vaccines (as seen in Table 1b) were significantly reduced in comparison to viral vector and non-replicating protein vaccinations, with vaccine efficacy seeing a marked improvement. In the last 2 decades, ≥50% (7/14) of drug-CDX co-developments failed to access the market at the same time.

The current process to identify and classify SARS-COV-2 variants is a detailed series of steps that takes up to a month from the time of a positive tests for COVID-19 to the identification and classification of a variant. Apparently, politics and government/state funding also contribute to the slow speed variants are identified.

### Research Practice Variations

In the analysis of research practice variations, there have been significant and unexplained variations between phases of co-development process time of companion diagnostics (Bartlett test P<0.001).

Similarly, there was a large amount of variation when looking at the process of COVID-19 vaccine development. This high level of variation is reflected in the high standard deviations that are seen in Table 1b, Table 2b, and table 3. There was variation in the number of days each phase of the clinical trial between the 6 different COVID-19 vaccines. While the transition times for both the non-replicating viral vector adjuvant and protein subunit matrix M vaccines ranged from 75 to 116 days between trials, the mRNA based Moderna and Pfizer vaccines averaged half that at only fifty-one days.

**Table 2a:**
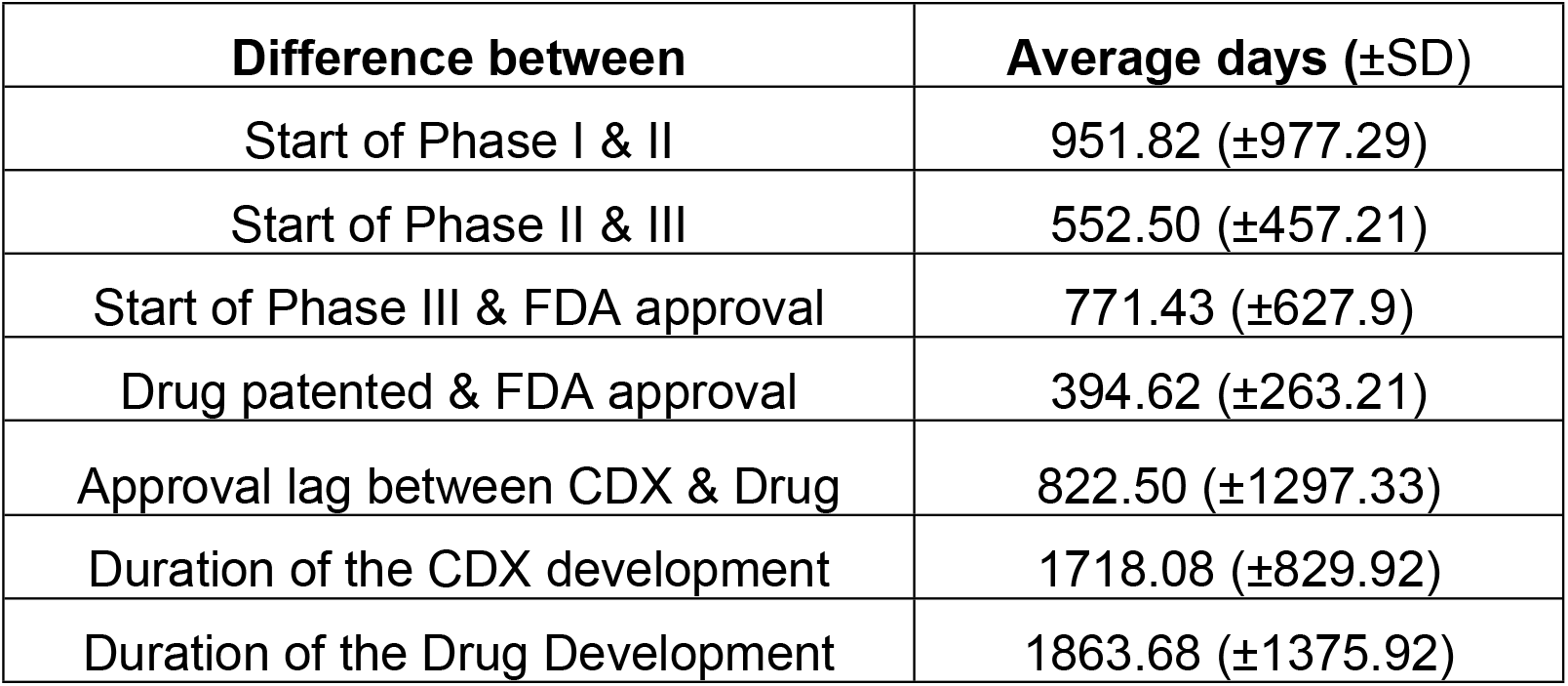
Difference between phases of co-development process and total development time of Companion Diagnostics

| **Difference between** | **Average days (±SD)** |
| --- | --- |
| Start of Phase I & II | 951.82 (±977.29) |
| Start of Phase II & III | 552.50 (±457.21) |
| Start of Phase III & FDA approval | 771.43 (±627.9) |
| Drug patented & FDA approval | 394.62 (±263.21) |
| Approval lag between CDX & Drug | 822.50 (±1297.33) |
| Duration of the CDX development | 1718.08 (±829.92) |
| Duration of the Drug Development | 1863.68 (±1375.92) |

**Table 2b:** Group specific average transition times (start of preclinical testing to FDA approval)

| Vaccine | Average | Standard Deviation |
| --- | --- | --- |
| mRNA | 207 | 123.04 |
| Non-replicating viral vector | 300 | 38.18 |
| Protein subunit + matrix M adjuvant | 465.50 | 232.64 |

**Table 3:** Subject participation in COVID-19 vaccine clinical trials

| **Development of Vaccine** | **Phase 1 subjects** | **Phase 2 subjects** | **Phase 3 subjects** |
| --- | --- | --- | --- |
| BioNTech/Pfizer | 200 | 200 | 44,000 |
| Moderna | 45 | 600 | 30,000 |
| AstraZeneca | 1,077 | 1,077 | 40,000 |
| Janssen (Johnson& Johnson) | 1,045 | 1,045 | 60,000 |
| Novavax | 131 | 1,500 | 33,000 |
| GSK/Sanofi | 440 | 722 | 5,414 |
| **Average (±SD)** | **489.7 (±461.8)** | **857.3 (±450.1)** | **35402 (±18079)** |

There was also large variation in participants in the drug trials of the vaccines investigated and between the different phases of drug trials. Table 3 shows the total subject numbers in vaccine trials were relatively similar between the mRNA-based, viral vector, and non-replicating protein vaccinations, with the Sanofi vaccine having the least participants at 6,576 subjects across all three phases. Other vaccines’ subject pools ranged from 30,645 for the Moderna to 62,090 for the Johnson & Johnson.

We also looked at the process of identifying and classifying variants to the SARS-COV-2 virus. Particular attention was given to the frequency of positive Covid-19 sequences submitted by 50 randomly selected countries which can be seen in Figure 2a. The frequency of positive Covid-19 sequences submitted by each of the 50 US states also shows enormous variation) Figure 2a).

**Figure 2a:**
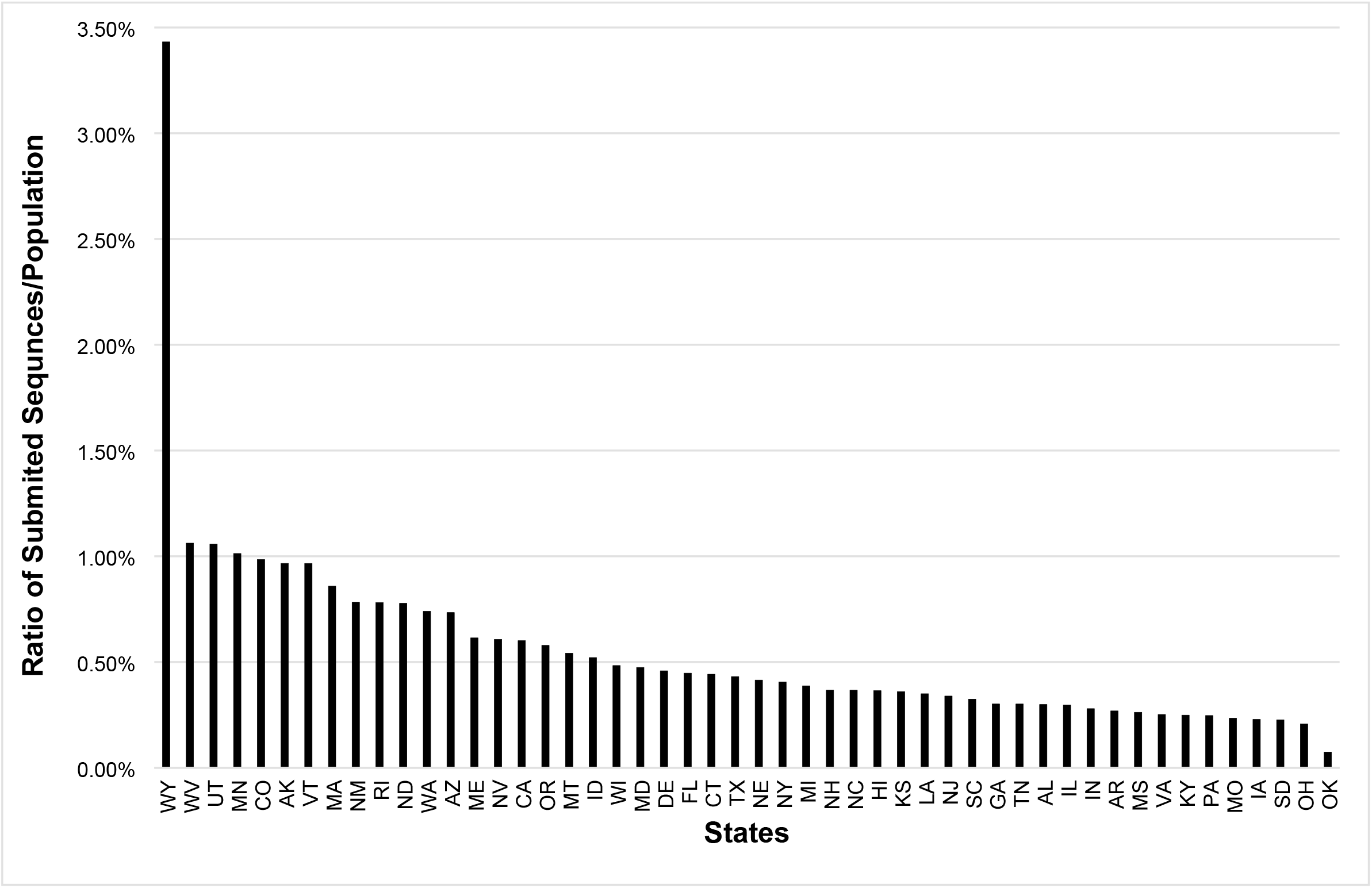
Ratio Sequences/ population of the 50 US States.

**Figure 2b:**
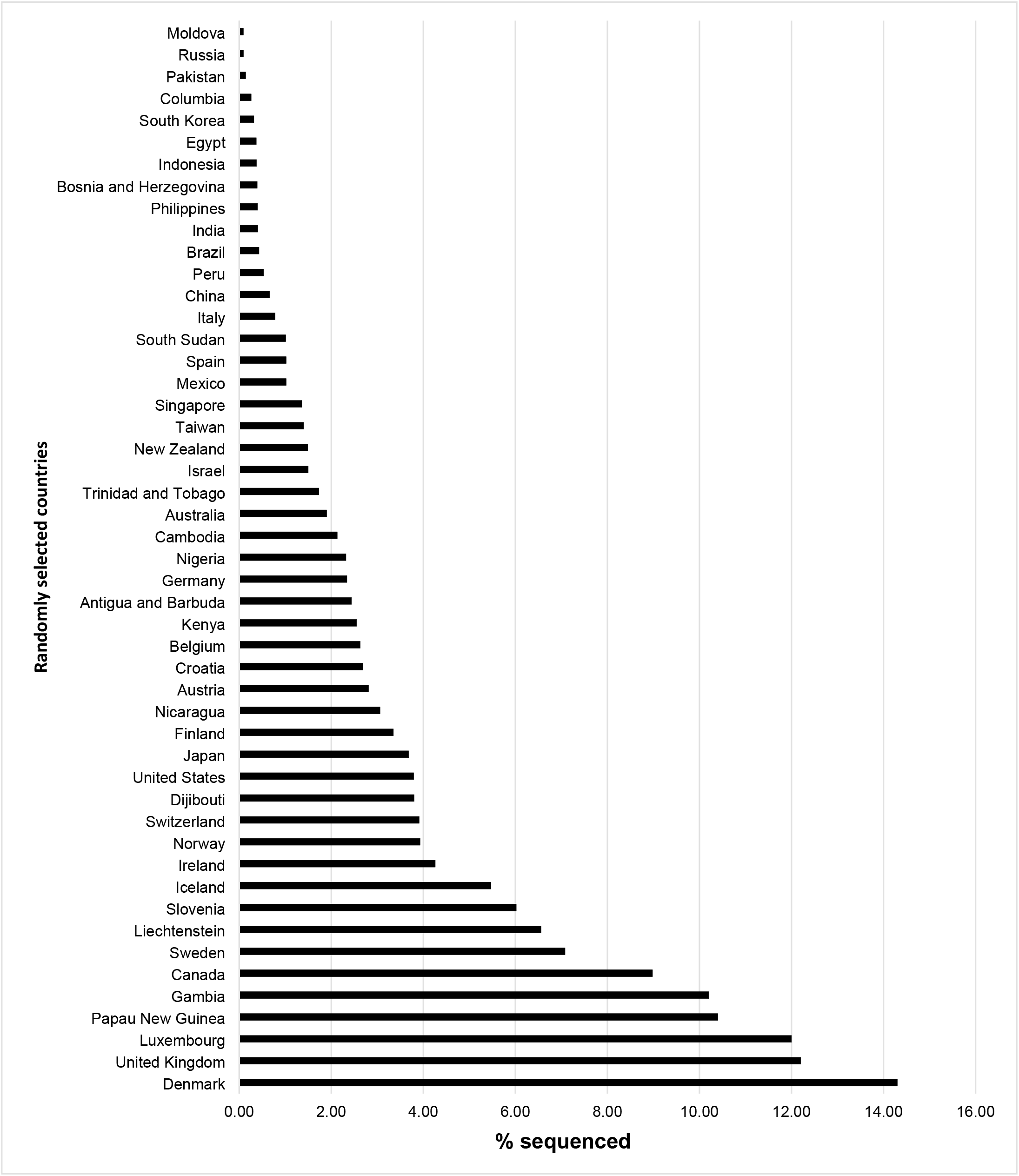
% Sequenced of Positive Covid-19 Samples of 50 Randomly Selected Countries.

Amongst the 50 randomly selected countries there was a high variability of the number of positive COVID-19 samples sequenced with an average 179,491 samples sequenced and an average percent sequenced of 3.28 with a standard deviation of 558600.04 and 3.55 respectively which can be seen in Table 2. Amongst all 50 US states the state with the highest ratio of positive samples sequenced per population was Wyoming with 3.4%. There seems to be a correlation between the population size of a country and the percentage sequenced. Iceland surprisingly had a higher percentage of samples sequenced.

## Discussion

Applications of our systematic research flow analysis methodology uncover many previously unexposed shortcomings. In vaccine development, COVID variant detection, and companion diagnostics development, we found major and largely unexplained variations of research processes. Such variations suggest that some of the excesses represent ineffective or wasteful use of resources like development time, certain level of patient recruitment in clinical trials or inconsistent sample collection for variant detection. The detected, surprisingly high rates of unexplained and unjustified variations in companion diagnostic development, vaccine development research and also in the identification of virus variants demand attention.

Our study defines an original, comprehensive methodology for the recognition of major deficiencies in research processes. Considering the unacceptably high ratio of non-reproducible research studies, our methodology is important for quality and reproducibility improvement in life sciences research. With the emergence of new electronic data sources and the pressing need for more trustworthy science, many research projects can be analyzed with our timely method. The propsed methodology is simple, structured and ready for wider use.

The increasing demand for precision medicine also indicates that the current co-development process needs a new paradigm. We suggest the early integration of drug and diagnostic components, which is crucial for fast and timely global commercialization and FDA approval simultaneously. The co-development process requires better understanding of gene biology of the disease and the drug mechanism of action to generate a strong biomarker hypothesis for prototype assay development and clinical trials. Simultaneously, the specificity of the companion diagnostic is also essential: a) for the true selection of patients who would be most likely to benefit for the specific drug, b) to recognize the patient at risk for the selected therapeutic product, and c) to monitor the response of the drug dosage, effectiveness, and safety of the selected therapy. The typical development of companion diagnostics needs to be contemporaneous.

It is noteworthy that the mRNA vaccine development times were significantly shorter than the protein and viral vector vaccine developments, two of which just gained FDA approval at the end of 2021 with another gaining approval in January 2022. There was a direct connection between mRNA research done over the last two decades to the research done on mRNA vaccines more recently, which drastically reduced production time and led to a record-setting COVID-19 vaccine development time.

In identification of COVID-19 variants, more worldwide cooperation would also benefit the identification process, particularly learning from the success of countries like Iceland and Denmark. The United States, despite having some of the most advanced technology, is behind many other countries regarding the number of samples sequenced. The time it takes from an increase in positive SARS-COV-2 cases to variant classification needs to be decreased. The frequency that sequences are submitted must increase in many but not all states. Future studies could look at how the political climate of a region affects the submission of sequences and the identification of disease variants as well as how governments affect variant research.

## Data Availability

Augusta University Commons

## Acknowledgement

The authors thank the faculty and students of the Clinical Laboratory Science Program and also team members of the Biomedical Research Innovation laboratory for helpful discussions. The authors also recognize partial funding support from the US National Institutes of Health (R01 GM146338-01).

## Abbreviations

R&I: Research and Innovation
COVID-19: coronavirus disease of 2019
SARS-COV-2: Severe acute respiratory syndrome coronavirus 2
PubMed: Publisher MEDLINE
USPTO: United States Patent and Trademark Office
R&D: Research and Development
ACE: Angiotensin Converting Enzyme
CDX: Companion Diagnostics

